# Parkinsonism in bipolar disorder: a clinical-neuroimaging study

**DOI:** 10.1101/2024.02.23.24303233

**Authors:** Yoshiyuki Nishio, Kiyomi Amemiya, Jun Ohyama

**Author notes:** **CORRESPONDENCE:** Yoshiyuki Nishio.

## Abstract

**Background:** Parkinsonism is a frequently encountered symptom in individuals with bipolar disorder (BD). It can be drug-induced, co-occurring with Parkinson’s disease (PD), or a genuine motor abnormality of BD itself. This study aims to clarify the primary pathophysiology of parkinsonism in BD.

**Methods:** Sixteen patients with BD and parkinsonism were recruited from consecutive patients who were referred to a neurology clinic at a tertiary psychiatric center. The patients underwent clinical assessments, dopamine transporter single-photon computed tomography (DAT-SPECT), cardiac MIBG scintigraphy, and morphometric MRI. The positivity or negativity of Lewy body disease (LBD) biomarkers was determined based on the visual assessment of DAT-SPECT and heart-to-mediastinum ratio on cardiac MIBG scintigraphy. Four out of the 16 participants received 300-600mg of levodopa.

**Results:** Thirteen patients were diagnosed with BD type 1, and 12 had experienced > 5 previous mood episodes. Parkinsonism developed more than 10 years after the onset of BD and after the age of 50 years in all patients. Four cases were positive for LBD biomarkers. Six patients with negative LBD biomarkers showed reduced striatal uptake with z-scores below -2.0. MRI morphometry revealed varying degrees of brain atrophy in most patients. Three of the 4 patients did not respond to 600mg of levodopa.

**Conclusions:** This study suggests that the majority of parkinsonism in BD is not due to PD/LBD. Parkinsonism may be a genuine motor abnormality of BD in late life.

## INTRODUCTION

Bipolar disorder (BD) is a modern psychiatric entity that has its roots in the 19th-century concept of manic-depressive insanity. This concept emphasized the cyclical and remitting course of the disorder and contrasted it with dementia praecox, a gradually progressive psychosis [1]. Subsequent research suggests that BD may have a neuroprogressive nature over the long term, in addition to being cyclical [2]. For example, BD is linked to an increased risk of developing dementia [3] and movement disorders [4] in later life.

Parkinsonism is a frequently encountered clinical feature in individuals with BD, often attributed to the adverse effects of medications. From the perspective of the emerging conceptualization of genuine motor abnormalities in psychiatric disorders [5], parkinsonism may be an intrinsic neuroprogressive feature of BD. In support of this, recent large cohort studies demonstrate that individuals with BD have an increased risk of developing Parkinson’s disease (PD) later in life [4]. However, it may be difficult to differentiate between PD and other forms of parkinsonism in these studies, as the diagnosis of PD was based solely on medical records [6].

Currently, the clinical features of parkinsonism observed in BD have not been fully described. Only a few studies have addressed the relationship between parkinsonism in BD and Lewy body disease (LBD) [6,7]. This study aimed to clarify these issues using multimodal neuroimaging techniques, including dopamine transporter single-photon emission computed tomography (DAT-SPECT) and cardiac 123I-metaiodo-benzylguanidine (MIBG) scintigraphy, as well as levodopa administration.

## METHODS

The study protocol adhered strictly to the tenets of the Declaration of Helsinki, Fourth Edition, and was approved by the Ethics Committee of Tokyo Metropolitan Matsuzawa Hospital. Written informed consent was obtained from all participants after a detailed explanation of the study.

### Participants

Participants were recruited from consecutive patients with BD who were referred to the neurology department of Tokyo Metropolitan Matsuzawa Hospital, the largest tertiary psychiatric hospital in Japan, between August 2019 and July 2022. All patients who have parkinsonism, which is defined as bradykinesia in combination with either rest tremor, rigidity, or both, were invited to participate in the study [8]. Sixteen patients who agreed to participate were included in the study.

### Clinical assessments

Motor symptoms were assessed using the Movement Disorder Society-Unified Parkinson’s Disease Rating Scale Part 3 (UPDRS) [9], the Unified Dystonia Rating Scale (UDRS) [10], the oculomotor part of the Progressive Nuclear Palsy Rating Scale (PNPRS) [11], the Abnormal Involuntary Movement Scale (AIMS) [12]. The Mini-mental State Examination (MMSE) was used to assess cognitive function [13]. All patients were euthymic and non-psychotic as assessed by Montgomery-Åsberg Depression Rating Scale (MADRS) [14], Young Mania Rating Scale (YMRS) [15], and the 6-item version of the Positive and Negative Syndrome Scale (PANSS) [16]. The motor features were characterized in detail using seven subscores of UPDRS part 3: speech (item 1), facial appearance (item 2), rigidity of the neck (a part of item 3), rigidity of the extremities (a part of item 3), movements of the extremities (items 4-8), gait/balance/speed (items 9-14), and tremor (items 15-18).

Profiles of motor symptoms (initial neurological symptoms, age of onset, and duration of motor symptoms), autonomic symptoms (bowel dysfunction and dysuria), and psychiatric disorders, number of mood episodes and hospitalizations, history of psychosis, catatonia, electroconvulsive therapy, suicide attempts, substance abuse, and sleep disturbance) were collected using a semi-structured questionnaire with reference to patient’s chart.

### Neuroimaging

#### DAT-SPECT

Patients were scanned on a dual-headed (INFINIA; GE Healthcare, MI, USA) or a triple-headed (GCA9300R; Canon Medical Systems, Tokyo, Japan) gamma camera 4h after intravenous injection of 167 MBq 123I-ioflupane. For analysis, 3.5-mm-thick axial images were reconstructed.

Three raters visually assessed blinded SPECT images according to the pre-defined grading [17]: Normal, largely symmetric tracer uptake in putamen and caudate nuclei; Grade 1, asymmetric uptake with normal/almost normal putamen uptake in one hemisphere and with a more marked reduction in the contralateral putamen; Grade 2, significant bilateral reduction in putamen uptake with activity confined to the caudate nuclei; Grade 3, virtually absent uptake bilaterally affecting both putamen and caudate nuclei. Grades 1-3 on one or more raters were considered abnormal.

A semi-quantitative analysis was performed using DaTView software (Nihon Medi-Pysics, Tokyo, Japan), in which the specific binding ratio (SBR) was defined as the mean counts of the entire striatum divided by the global brain [18]. Age-corrected z-scores of SBR were calculated based on the Japanese multicenter database of healthy controls [19].

#### Cardiac MIBG scintigraphy

Data were collected using the SPECT cameras described above after 20 min (early image) and 240 min (delayed image) after intravenous injection of 111 MBq of 123I-MIBG. Regions of interest were drawn around the whole heart (H) and mediastinum (M). The H/M ratio was calculated from the average number of counts per pixel in the H and M. The H/M ratio on early or delayed images ≤ 2.0 was considered abnormal.

#### Definition of the positivity for the LBD biomarker

A decrease in DAT activity is a finding observed in various neurological and psychiatric conditions and is not specific to LBD [20,21]. When diagnosing PD, it is necessary to consider intrastriatal topography, such as the reduction in the posterior putamen. In this regard, visual assessment outperforms SBR [22]. A decreased H/M ratio in MIBG myocardial scintigraphy is highly specific to LBD. Taking all these factors into account, our study classified cases with abnormal visual assessment of DAT-SPECT or decreased H/M ratio as positive for the LBD biomarker.

#### Magnetic resonance imaging

3D MPRAGE images (TR, 1800ms; TE, 2.92ms; FA 10deg; FOV, 240; Matrix size, 256; Slice thickness 1mm) were acquired on a 3T MRI scanner (MAGNETOM Skyra; Erlangen, Germany). Voxel-based morphometry analysis was performed using SPM12 (https://www.fil.ion.ucl.ac.uk/spm/software/spm12/) and BAAD

(https://www.shiga-med.ac.jp/hqbioph/iBaad/page0.html). The BAAD quantitatively calculates the extent of gray matter volume reduction based on a comparison with MR images from the IXI database of age-matched healthy controls (Information Extraction from Images, Control Group IXI60 and IXI7080). Total intracranial volume, age, and sex were included as covariates. Voxels with a z-score of >2 with a cluster extent > 50 voxels were displayed on the MNI Average Brain. Whole brain volume was calculated as [(“total gray matter volume” + “total white matter volume”)/”total intracranial volume”]*1000.

### Levodopa administration

All participants received a comprehensive explanation of the potential benefits and adverse effects of levodopa. Four patients consented to taking the medication. This part of the study was preregistered in UMIN-CTR (University Hospital Medical Information Network Clinical Research Registration System) as UMIN000038397.

Levodopa/carbidopa was started at a dose of 100 mg twice daily and gradually increased to 100-200 mg three times daily over 2 months. The maximum daily dose was set at 600 mg. Baseline and post-administration assessments were conducted less than 1 month before levodopa initiation and 3-6 months after the baseline assessment, respectively.

## RESULTS

### Clinical profiles

Demographic data and clinical profiles of patients are shown in **Tables 1, 2, S1, and S2**. Of the 16 patients, 13 had BD type 1, and 12 had 5 or more previous mood episodes. Overall, the disease severity of BD was high in the present cases. All patients developed parkinsonism more than 10 years after the onset of BD and after the age of 50 years; none had a history of catatonia or electroconvulsive therapy. Eight patients were taking lithium and 12 patients were taking valproic acid or carbamazepine. Antipsychotic drugs that the participants took include quetiapine (4 cases), aripiprazole (4), olanzapine (3), and risperidone (1). The mean defined daily doses for antipsychotic drugs/olanzapine equivalent dose (DDD-OLA) was 7.5±8.3 mg. The features of motor symptoms were indistinguishable from those of idiopathic PD.

**Table 1.**
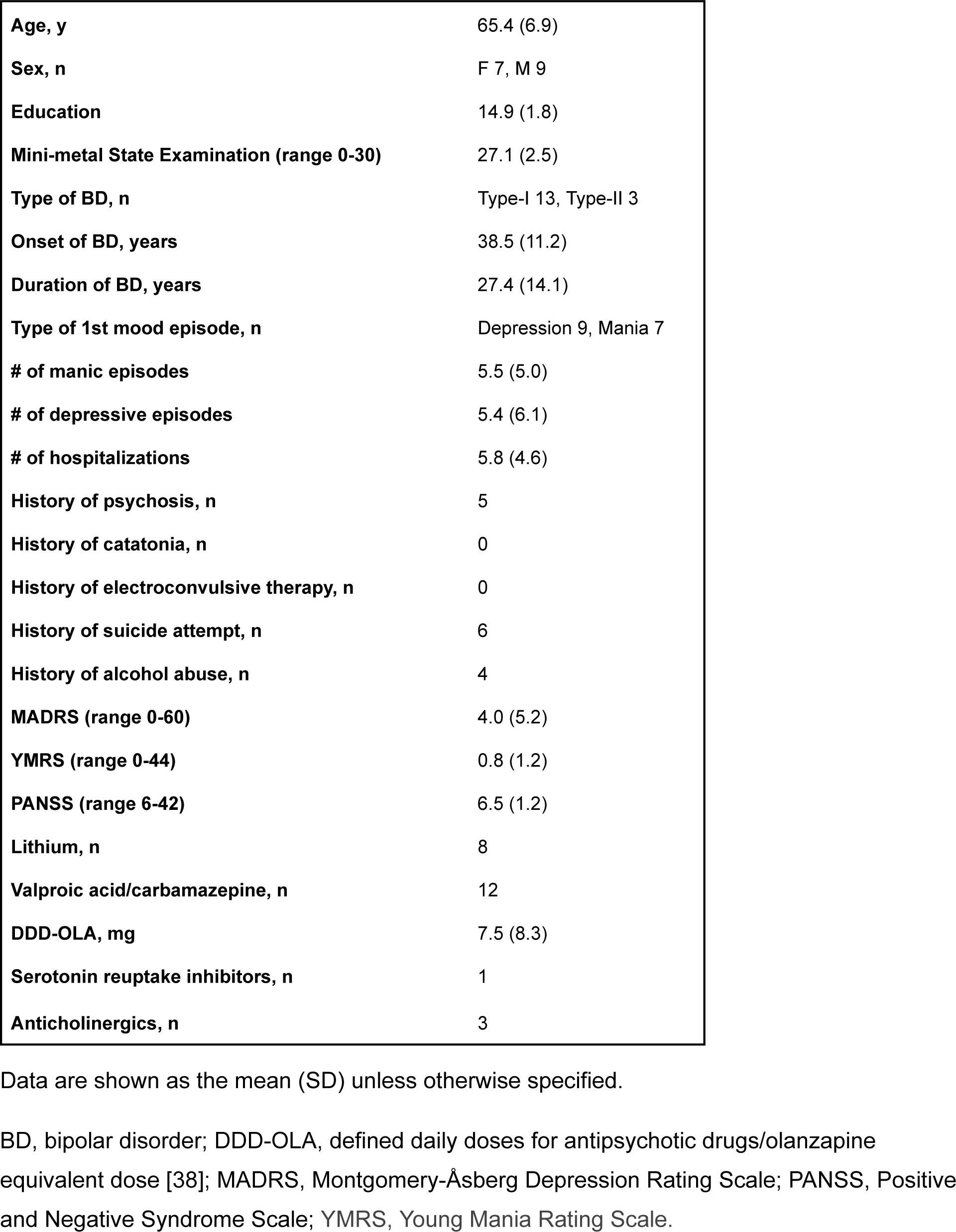
Demographic data and clinical characteristics of patients.

**Table 2.**
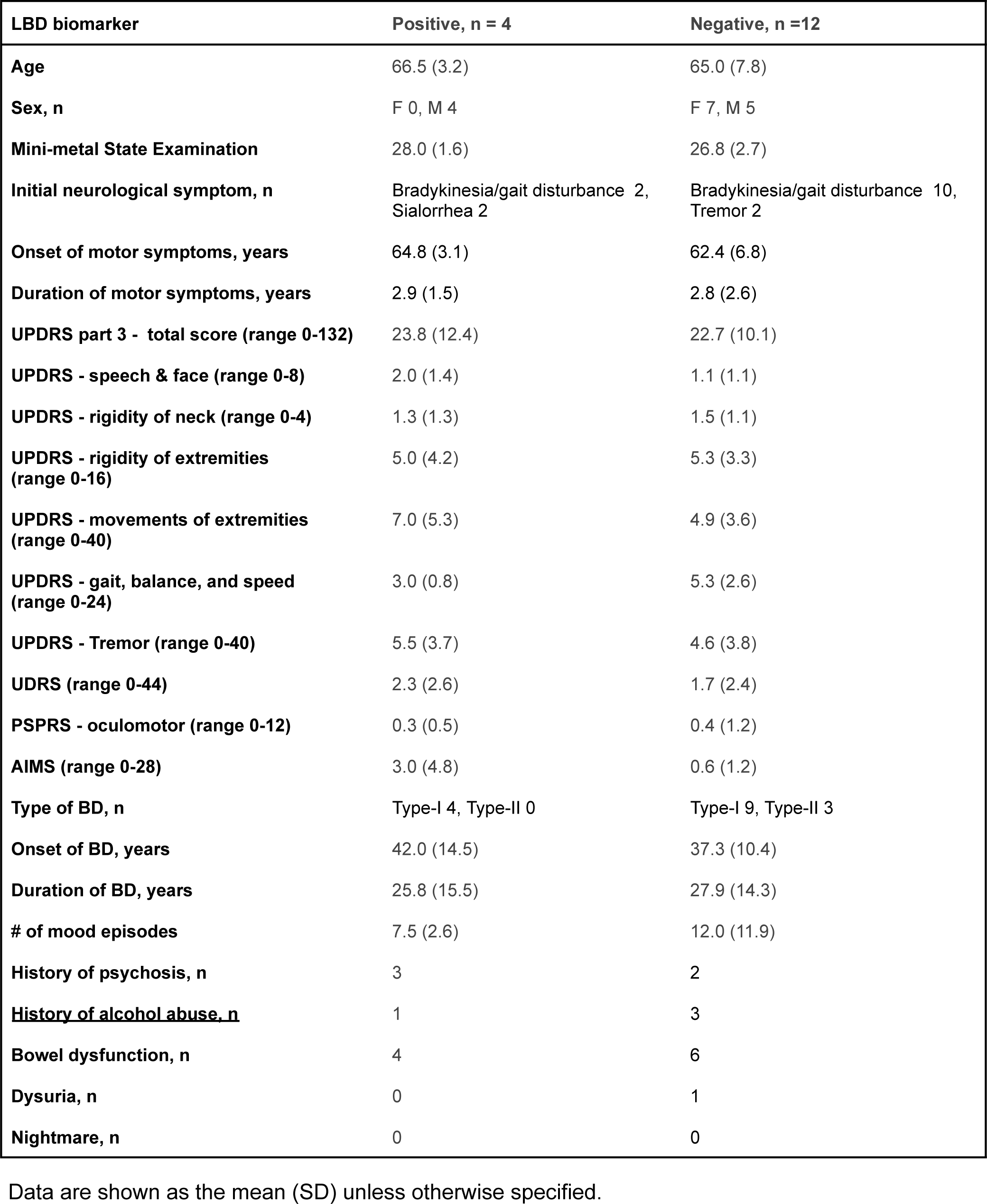
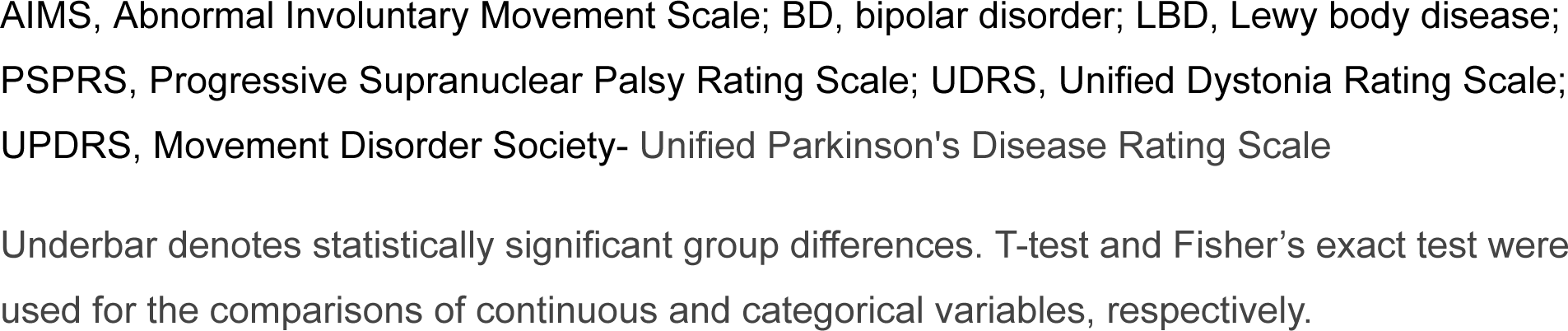
Clinical profiles of biomarker-positive and -negative patients.

| LBD biomarker | Positive, n = 4 | Negative, n =12 |
| --- | --- | --- |
| Age | 66.5 (3.2) | 65.0 (7.8) |
| Sex, n | F 0, M 4 | F 7, M 5 |
| Mini-metal State Examination | 28.0 (1.6) | 26.8 (2.7) |
| Initial neurological symptom, n | Bradykinesia/gait disturbance 2,<br>Sialorrhea 2 | Bradykinesia/gait disturbance 10,<br>Tremor 2 |
| Onset of motor symptoms, years | 64.8 (3.1) | 62.4 (6.8) |
| Duration of motor symptoms, years | 2.9 (1.5) | 2.8 (2.6) |
| UPDRS part 3 - total score (range 0-132) | 23.8 (12.4) | 22.7 (10.1) |
| UPDRS - speech & face (range 0-8) | 2.0 (1.4) | 1.1 (1.1) |
| UPDRS - rigidity of neck (range 0-4) | 1.3 (1.3) | 1.5 (1.1) |
| UPDRS - rigidity of extremities<br>(range 0-16) | 5.0 (4.2) | 5.3 (3.3) |
| UPDRS - movements of extremities<br>(range 0-40) | 7.0 (5.3) | 4.9 (3.6) |
| UPDRS - gait, balance, and speed<br>(range 0-24) | 3.0 (0.8) | 5.3 (2.6) |
| UPDRS - Tremor (range 0-40) | 5.5 (3.7) | 4.6 (3.8) |
| UDRS (range 0-44) | 2.3 (2.6) | 1.7 (2.4) |
| PSPRS - oculomotor (range 0-12) | 0.3 (0.5) | 0.4 (1.2) |
| AIMS (range 0-28) | 3.0 (4.8) | 0.6 (1.2) |
| Type of BD, n | Type-I 4, Type-II 0 | Type-I 9, Type-II 3 |
| Onset of BD, years | 42.0 (14.5) | 37.3 (10.4) |
| Duration of BD, years | 25.8 (15.5) | 27.9 (14.3) |
| # of mood episodes | 7.5 (2.6) | 12.0 (11.9) |
| History of psychosis, n | 3 | 2 |
| <u>History of alcohol abuse, n</u> | 1 | 3 |
| Bowel dysfunction, n | 4 | 6 |
| Dysuria, n | 0 | 1 |
| Nightmare, n | 0 | 0 |

Data are shown as the mean (SD) unless otherwise specified.
AIMS, Abnormal Involuntary Movement Scale; BD, bipolar disorder; LBD, Lewy body disease; PSPRS, Progressive Supranuclear Palsy Rating Scale; UDRS, Unified Dystonia Rating Scale; UPDRS, Movement Disorder Society- Unified Parkinson's Disease Rating Scale
Underbar denotes statistically significant group differences. T-test and Fisher's exact test were used for the comparisons of continuous and categorical variables, respectively.

In comparisons between patient groups positive (n = 4) and negative (n = 12) for LBD biomarkers, those negative for the biomarkers were more likely to have a history of alcohol abuse than those positive for the biomarkers. The groups did not differ significantly in terms of other variables, such as movement disorder characteristics, duration of BD, number of mood episodes, and autonomic symptoms.

### Neuroimaging

Four cases were positive for LBD biomarkers. Of these cases, 2 were abnormal on DAT-SPECT (both grade 1), 1 was abnormal on MIBG, and 1 was abnormal on both DAT-SPECT (grade 3) and MIBG (**Figures 1 and S1; Tables 3 and S3**). All 12 LBD biomarker-negative patients had a below-normal-mean SBR with a z-score < 0. In 6 of these patients, the SBR z-score was below -2.0. MRI voxel-based morphometry revealed varying degrees of brain atrophy in most patients, primarily located in the frontal lobes (**Figures 1 and S2**). There was no significant difference in whole brain volumes between the biomarker-positive and negative patient groups (**Table 3**).

**Figure 1.**
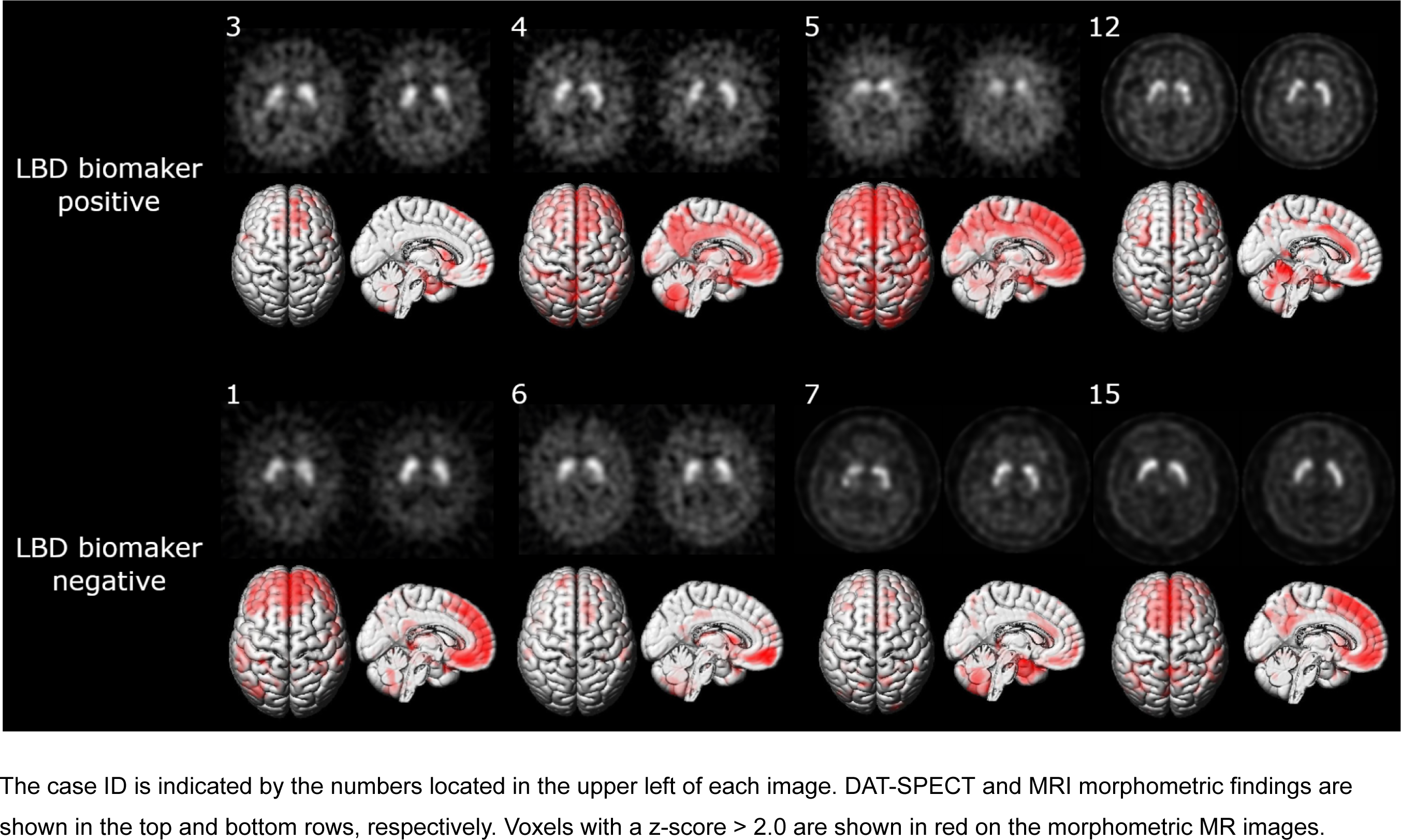
Neuroimaging findings of patients with positive and negative Lewy body biomarkers. The case ID is indicated by the numbers located in the upper left of each image. DAT-SPECT and MRI morphometric findings are shown in the top and bottom rows, respectively. Voxels with a z-score > 2.0 are shown in red on the morphometric MR images.

**Table 3.**
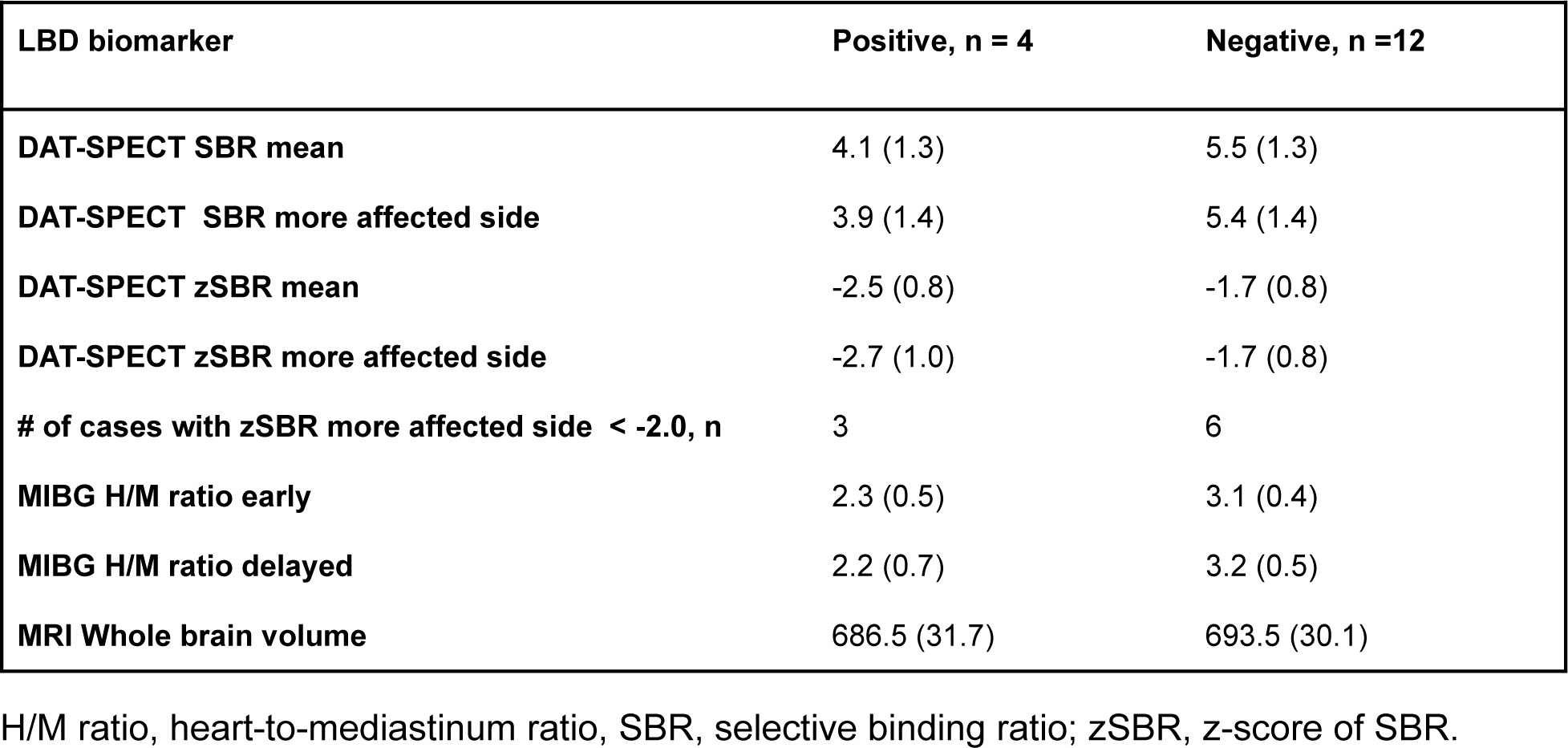
Neuroimaging results.

### Brain-behavior correlations

For exploratory purposes, we conducted correlation analyses of the UPDRS part 3 total score with brain imaging or clinical variables (Pearson’s correlations) (**Figure S3**). No significant correlations were found in all of the analyses. Although statistically insignificant, a scatter plot indicates a trend of correlation between the total scores of UPDRS part 3 and age-corrected z-scores of SBR.

### Effects of levodopa

Of the 4 cases, motor symptoms were improved in only 1 case, as assessed by the UPDRS part 3 total score. This case had an abnormal pre-treatment DAT-SPECT (**Table 4**). However, his motor symptoms did not worsen when levodopa was discontinued 2 years later, suggesting that the observed improvement may not be due to levodopa treatment. Levodopa was well tolerated in all cases, and no worsening of psychiatric symptoms as assessed by MADRS, YMRS, and PANSS was observed.

**Table 4.** Changes in motor and psychiatric symptoms before and after receiving L-dopa.

| Pt | DAT /MIBG | L-dopa (mg) | UPDRS part 3 total | UPDRS speech & face | UPDRS rigidity neck | UPDRS rigidity extremities | UPDRS movement extremities | UPDRS balance & speed | UPDRS tremor | UDRS | PSPRS oculo-motor | AIMS | MADRS | YMRS | PANSS |
| --- | --- | --- | --- | --- | --- | --- | --- | --- | --- | --- | --- | --- | --- | --- | --- |
| 2 | - / - | 600 | 35/48 | 4/4 | 3/3 | 10/11 | 8/15 | 6/13 | 4/2 | 1.5/3.5 | 4/4 | 0/0 | 0/0 | 4/0 | 6/6 |
| 3 | + / - | 600 | 18/29 | 3/3 | 1/3 | 6/7 | 4/6 | 3/9 | 1/1 | 0/0 | 1/3 | 0/0 | 5/0 | 0/0 | 6/6 |
| 4 | + / - | 300 | **40/23** | 0/0 | 3/2 | 10/4 | 14/8 | 4/3 | 9/6 | 5/0 | 0/0 | 2/2 | 15/15 | 2/2 | 6/6 |
| 7 | - / - | 600 | 20/12 | 1/0 | 2/0 | 5/2 | 2/2 | 6/6 | 4/2 | 2.5/2.5 | 0/0 | 0/0 | 0/16 | 0/2 | 6/6 |

Data are presented as pre/post. DAT-SPECT (DAT) was considered abnormal if the visual assessment was in grades 1-3. Cardiac MIBG scintigraphy was considered abnormal if the early or delayed H/M ratio was < 2.2.
AIMS, Abnormal Involuntary Movement Scale; MADRS, Montgomery-Åsberg Depression Rating Scale; PANSS, Positive and Negative Syndrome Scale; PSPRS, Progressive Supranuclear Palsy Rating Scale; UDRS, Unified Dystonia Rating Scale UPDRS, Movement Disorder Society- Unified Parkinson's Disease Rating Scale; YMRS, Young Mania Rating Scale

## DISCUSSION

### Is parkinsonism in BD caused by PD/LBD?

In previous studies of patients with a confirmed diagnosis of PD at longitudinal follow-up, the sensitivity of DAT-SPECT by visual assessment at baseline was reported to be >90%, with false negative results <5% [23,24]. Compared to these studies, the rate of DAT-SPECT abnormalities in the present study is considerably low, i.e. 18.8% of all participants (3 of 16 patients). Furthermore, the proportion of patients responding to levodopa was lower than that usually observed in PD, with 3 of the 4 patients failing to respond to 600 mg of levodopa. These findings suggest that the majority of parkinsonism in BD may not be due to PD/LBD. It should be noted that striatal uptake was decreased on semiquantitative assessment (SBR z-score < -2.0) in 6 of 13 cases (46.1%) that were considered normal on visual assessment. A previous study of the early PD cohort reported that 80% of cases judged normal on visual assessment had decreased striatal uptake on semiquantitative assessment, and a small percentage of patients with such a discrepancy may be diagnosed with PD at longitudinal follow-up [24,25]. During long-term follow-up, some patients who were considered normal for the LBD biomarker in the present study may have been rediagnosed with PD.

MIBG is a highly specific biomarker for LBD with few abnormal findings in other neurodegenerative diseases. It is also highly sensitive for early PD, with a sensitivity exceeding 80% when the H/M ratio of 1.5-2.0 is used as a cut-off for patients with Hoehn and Yahr stages 1.5-2.5 [26]. In the present study, only 2 out of 16 cases (12.5%) were positive even when using a lenient H/M ratio cut-off of 2.0. Again, this finding suggests that parkinsonism is not commonly associated with PD/LBD in patients with BD.

### Is parkinsonism in BD drug-induced?

Patients with BD are often prescribed antidopaminergic antipsychotics, which may contribute to the development of motor symptoms. According to a recent meta-analysis, extrapyramidal symptoms are dose-dependent for most antipsychotic drugs. Extrapyramidal symptoms increase significantly when D2 receptor occupancy exceeds 75-85%, with the exception of quetiapine, which carries no risk of extrapyramidal symptoms at any dose [27]. Previous studies using DAT-SPECT did not find significant effects of antipsychotics on striatal uptake [28]. In the present study, we did not observe significant effects of antipsychotic doses on motor symptom severity or SBR on DAT-SPECT (**Figure S3**). Our findings may be related to the low doses of antipsychotics with low D2 receptor affinity taken by most of the participants.

Lithium is known to increase the risk of parkinsonism and has been shown to affect DAT activity in the striatum [29,30]. In the present study, there was no significant difference in the severity of motor impairment or SBR on DAT-SPECT between cases taking lithium and those not taking lithium (**Figure S3b**). Antidepressants such as mirtazapine and mianserin, which act on α2-adrenergic receptors, are known to significantly affect DAT-SPECT. However, none of the participants in the present study were receiving these drugs [28].

### Genuine motor abnormalities, neuroprogression, and parkinsonism in BD

Drug-induced parkinsonism is the most common clinical diagnosis for parkinsonism in BD [6]. The diagnosis of drug-induced parkinsonism is based solely on the temporal relationship between the onset of parkinsonism and the start of the suspected drug. In research practice, normal findings on DAT-SPECT are additionally used to exclude complications of PD/LBD. However, in addition to PD/LBD and drug-induced parkinsonism, it is important to consider the possibility of genuine motor abnormalities or neuroprogressive symptoms of BD itself [2,5].

Neurological soft signs, a collection of subtle neurological abnormalities, are frequently observed in individuals with BD [31]. Additionally, patients with bipolar disorder (BD) are reportedly more susceptible to drug-induced parkinsonism than patients with schizophrenia [32].

Recent neuroimaging studies have shown that patients with BD exhibit atrophy in various brain regions, including cortical and subcortical structures [33]. The degree of structural changes increases with age and is more pronounced in patients with type 1 BD or frequent manic episodes [33,34]. Consistent with these findings, the majority of participants in the present study had type 1 BD, a disease duration of over 10 years, and exhibited brain atrophy in various regions. It is unclear from these findings whether there is a cumulative negative impact of manic episodes on brain structure. Instead, neuroprogression may not be a general rule in BD but may occur in a specific subtype of BD [35].

The dopamine hypothesis of bipolar disorder (BD) has been proposed based on the similarity between behavioral changes induced by amphetamine administration and manic symptoms, as well as the effects of antidopaminergic medications on manic symptoms. Although dopaminergic neuroimaging studies have revealed various abnormalities in BD, there are also many inconsistencies in the findings across studies [36,37]. Some studies have reported that striatal dopamine transporter (DAT) activity is decreased only in the manic state of BD [36,37], while others have reported a decrease also in euthymia [20]. The present study found that most BD patients with parkinsonism had reduced striatal DAT activity, and there was a trend of correlation between striatal DAT activity and the severity of parkinsonism. This finding suggests that striatal DAT activity may be linked to chronic clinical features, including parkinsonism, in addition to mood states. It is important to note that all of these molecular neuroimaging studies, including ours, have been small cross-sectional studies. The variability of the results may be due to sampling bias reflecting the clinical heterogeneity of BD. A larger molecular neuroimaging study is necessary, which considers clinical variables beyond mood states.

### Limitations

The main limitations of this study are the small sample size and recruitment from a single neurology clinic. These factors may have biased the participants towards BD with relatively high motor severity. Inference of the underlying pathophysiology of parkinsonism from neuroimaging findings should also be based on a larger sample size of patients with a wider range of severity. The present findings are not immediately generalizable and should be considered a pilot for larger studies on the pathogenesis of parkinsonism as a late manifestation of BD.

## Data Availability

All data produced in the present work are contained in the manuscript

## Supplementary materials

**Table S1.** Demographic data and motor symptoms of patients.

| Pt | Age<br>Sex | Edu | MMSE | Onset<br>motor<br>(y.o.) | Duration<br>motor<br>(y) | Initial<br>motor<br>symptom | UPDRS<br>part III<br>total<br>/132 | UPDRS<br>speech<br>& face<br>/8 | UPDRS<br>rigidity<br>neck<br>/4 | UPDRS<br>rigidity<br>UE & LE<br>/16 | UPDRS<br>Move-<br>ment<br>UE & LE<br>/40 | UPDRS<br>gait,<br>balance<br>& speed<br>/24 | UPDRS<br>Tremor<br>/40 | UDRS<br>/44 | PSPRS<br>oculo-<br>motor<br>/12 | AIMS<br>/28 |
| --- | --- | --- | --- | --- | --- | --- | --- | --- | --- | --- | --- | --- | --- | --- | --- | --- |
| 1 | 60'sM | 16 | 29 | 60's | 3 | Brady<br>-kinesia | 37 | 3 | 3 | 11 | 14 | 6 | 6 | 6 | 0 | 3 |
| 2 | 60'sM | 12 | 26 | 60's | 3 | Brady<br>-kinesia | 35 | 8 | 3 | 10 | 12 | 6 | 4 | 2 | 4 | 0 |
| 3 | 60'sM | 16 | 26 | 60's | 2 | Gait | 18 | 5 | 1 | 6 | 6 | 3 | 1 | 0 | 1 | 0 |
| 4 | 60'sM | 16 | 27 | 60's | 3 | Gait | 40 | 0 | 3 | 10 | 22 | 4 | 9 | 5 | 0 | 2 |
| 5 | 70'sM | 16 | 30 | 60's | 5 | Sialorrhea | 11 | 3 | 0 | 0 | 2 | 2 | 4 | 0 | 0 | 0 |
| 6 | 70'sF | 14 | 22 | 70's | 1 | Gait | 20 | 3 | 2 | 5 | 3 | 6 | 4 | 3 | 0 | 0 |
| 7 | 60'sM | 16 | 27 | 60's | 5 | Tremor | 13 | 4 | 0 | 1 | 3 | 3 | 4 | 0 | 0 | 2 |
| 8 | 60'sF | 16 | 28 | 60's | 1 | Gait | 32 | 4 | 2 | 4 | 3 | 6 | 15 | 0 | 0 | 0 |
| 9 | 50'sM | 18 | 30 | 50's | 1 | Brady<br>-kinesia | 10 | 3 | 0 | 2 | 4 | 5 | 0 | 0 | 0 | 0 |
| 10 | 60'sF | 16 | 30 | 60's | 3 | Gait,<br>Clumsiness | 24 | 1 | 2 | 8 | 10 | 3 | 4 | 0 | 0 | 0 |
| 11 | 50'sF | 14 | 24 | 50's | 1 | Gait | 17 | 0 | 1 | 4 | 2 | 3 | 7 | 0 | 1 | 0 |
| 12 | 60'sM | 12 | 29 | 60's | 1.5 | Sialorrhea | 26 | 4 | 1 | 4 | 11 | 3 | 8 | 4 | 0 | 10 |
| 13 | 50'sM | 16 | 26 | 60's | 10 | Gait | 17 | 1 | 1 | 2 | 8 | 8 | 1 | 5 | 0 | 0 |
| 14 | 50'sF | 14 | 29 | 50's | 1 | Brady | 19 | 0 | 2 | 5 | 8 | 6 | 2 | 0 | 0 | 0 |
|  |  |  |  |  |  | -kinesia |  |  |  |  |  |  |  |  |  |  |
| 15 | 50'sF | 14 | 28 | 50's | 2 | Temor | 11 | 0 | 0 | 3 | 4 | 1 | 5 | 0 | 0 | 3 |
| 16 | 70'sF | 12 | 23 | 70's | 2 | Brady<br>-kinesia | 37 | 0 | 2 | 8 | 15 | 11 | 3 | 6 | 0 | 0 |

**Table S2.**
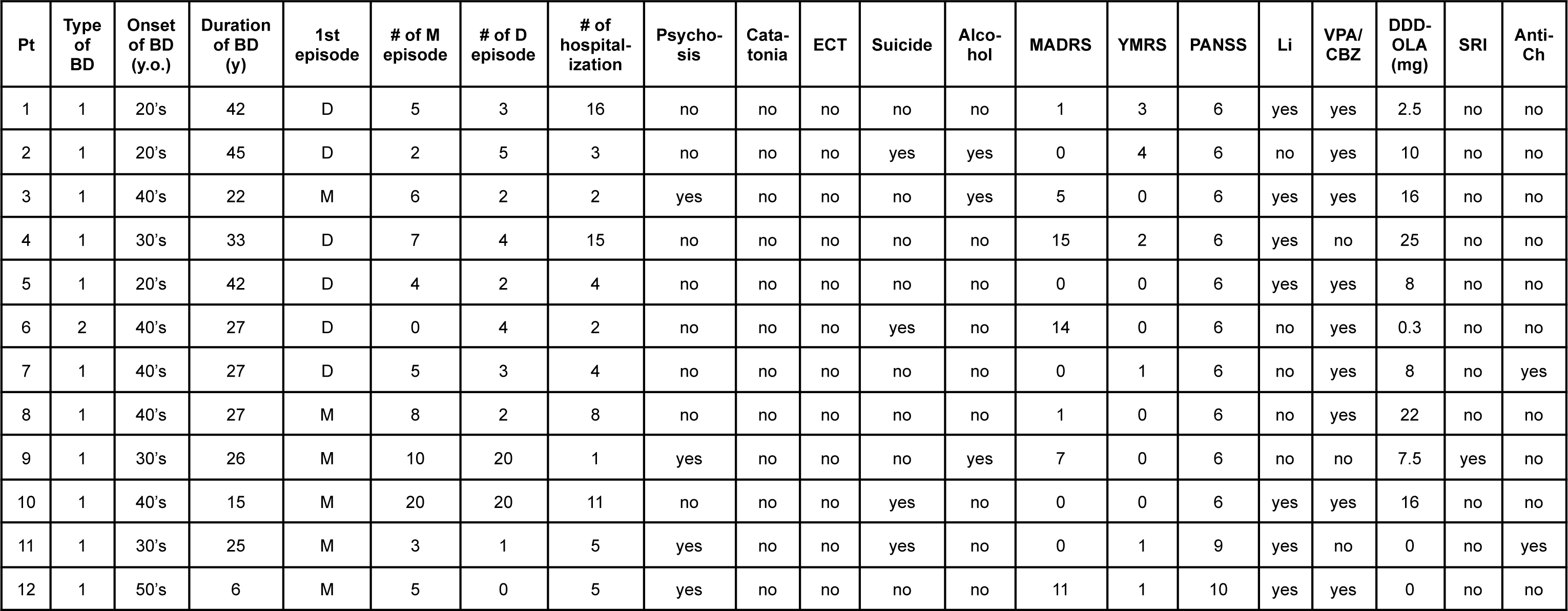
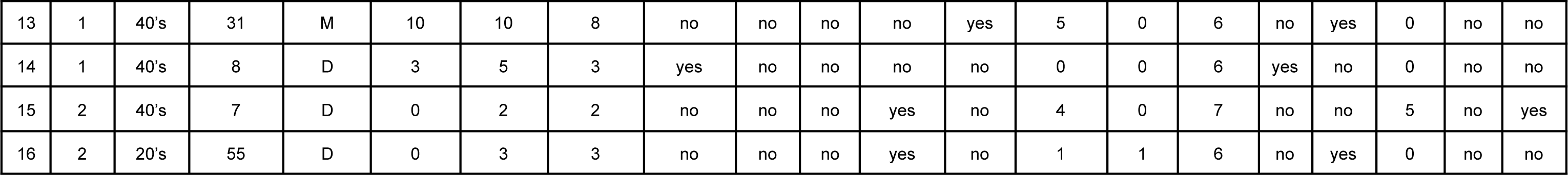
Psychiatric backgrounds of patients.

| Pt | Type of BD | Onset of BD (y.o.) | Duration of BD (y) | 1st episode | # of M episode | # of D episode | # of hospital-ization | Psycho-sis | Cata-tonia | ECT | Suicide | Alco-hol | MADRS | YMRS | PANSS | Li | VPA/CBZ | DDD-OLA (mg) | SRI | Anti-Ch |
| --- | --- | --- | --- | --- | --- | --- | --- | --- | --- | --- | --- | --- | --- | --- | --- | --- | --- | --- | --- | --- |
| 1 | 1 | 20's | 42 | D | 5 | 3 | 16 | no | no | no | no | no | 1 | 3 | 6 | yes | yes | 2.5 | no | no |
| 2 | 1 | 20's | 45 | D | 2 | 5 | 3 | no | no | no | yes | yes | 0 | 4 | 6 | no | yes | 10 | no | no |
| 3 | 1 | 40's | 22 | M | 6 | 2 | 2 | yes | no | no | no | yes | 5 | 0 | 6 | yes | yes | 16 | no | no |
| 4 | 1 | 30's | 33 | D | 7 | 4 | 15 | no | no | no | no | no | 15 | 2 | 6 | yes | no | 25 | no | no |
| 5 | 1 | 20's | 42 | D | 4 | 2 | 4 | no | no | no | no | no | 0 | 0 | 6 | yes | yes | 8 | no | no |
| 6 | 2 | 40's | 27 | D | 0 | 4 | 2 | no | no | no | yes | no | 14 | 0 | 6 | no | yes | 0.3 | no | no |
| 7 | 1 | 40's | 27 | D | 5 | 3 | 4 | no | no | no | no | no | 0 | 1 | 6 | no | yes | 8 | no | yes |
| 8 | 1 | 40's | 27 | M | 8 | 2 | 8 | no | no | no | no | no | 1 | 0 | 6 | no | yes | 22 | no | no |
| 9 | 1 | 30's | 26 | M | 10 | 20 | 1 | yes | no | no | no | yes | 7 | 0 | 6 | no | no | 7.5 | yes | no |
| 10 | 1 | 40's | 15 | M | 20 | 20 | 11 | no | no | no | yes | no | 0 | 0 | 6 | yes | yes | 16 | no | no |
| 11 | 1 | 30's | 25 | M | 3 | 1 | 5 | yes | no | no | yes | no | 0 | 1 | 9 | yes | no | 0 | no | yes |
| 12 | 1 | 50's | 6 | M | 5 | 0 | 5 | yes | no | no | no | no | 11 | 1 | 10 | yes | yes | 0 | no | no |
| 13 | 1 | 40's | 31 | M | 10 | 10 | 8 | no | no | no | no | yes | 5 | 0 | 6 | no | yes | 0 | no | no |
| 14 | 1 | 40's | 8 | D | 3 | 5 | 3 | yes | no | no | no | no | 0 | 0 | 6 | yes | no | 0 | no | no |
| 15 | 2 | 40's | 7 | D | 0 | 2 | 2 | no | no | no | yes | no | 4 | 0 | 7 | no | no | 5 | no | yes |
| 16 | 2 | 20's | 55 | D | 0 | 3 | 3 | no | no | no | yes | no | 1 | 1 | 6 | no | yes | 0 | no | no |

**Table S3.** Neuroimaging results.

| Pt | SBR mean | SBR more affected side | SBR less affected side | zSBR mean | zSBR more affected side | zSBR less affected side | DAT visual rating | MIBG H/M early | MIBG H/M delayed | WBV |
| --- | --- | --- | --- | --- | --- | --- | --- | --- | --- | --- |
| 1 | 7.0 | 6.8 | 7.1 | -0.3 | -0.4 | -0.2 | 0/0/0 | 3.2 | 3.4 | 663.6 |
| 2 | 5.8 | 5.6 | 5.9 | -1.3 | -1.4 | -1.2 | 0/0/0 | 4.0 | 4.2 | 701.6 |
| **<u>3</u>** | 3.6 | 3.5 | 3.8 | -2.8 | -2.9 | -2.7 | **<u>0/1/0</u>** | 3.0 | 3.1 | 692.0 |
| **<u>4</u>** | 4.7 | 4.5 | 5.0 | -2.2 | -2.4 | -2.0 | **<u>1/1/1</u>** | 2.3 | 2.3 | 640.3 |
| **<u>5</u>** | 2.4 | 2.1 | 2.8 | -3.6 | -3.9 | -3.4 | **<u>2/2/2</u>** | 2.2 | **<u>2.0</u>** | 704.8 |
| 6 | 4.7 | 4.6 | 4.8 | -1.9 | -2.0 | -1.9 | 0/0/0 | 2.9 | 2.4 | 695.3 |
| 7 | 4.9 | 4.8 | 5.0 | -1.9 | -1.9 | -1.8 | 0/0/0 | 3.4 | 3.8 | 671.2 |
| 8 | 7.0 | 6.9 | 7.0 | -0.4 | -0.5 | -0.4 | 0/0/0 | 3.0 | 2.9 | 723.8 |
| 9 | 4.8 | 4.8 | 4.9 | -2.3 | -2.3 | -2.2 | 0/0/0 | 2.8 | 2.7 | 727.7 |
| 10 | 6.7 | 6.4 | 7.1 | -1.1 | -1.3 | -0.8 | 0/0/0 | 2.8 | 3.0 | 742.0 |
| 11 | 5.5 | 5.5 | 5.6 | -2.2 | -2.2 | -2.1 | 0/0/0 | 3.1 | 3.3 | 683.8 |
| **<u>12</u>** | 5.4 | 5.4 | 5.5 | -1.6 | -1.6 | -1.5 | 0/0/0 | **<u>1.6</u>** | **<u>1.5</u>** | 709.0 |
| 13 | 3.0 | 2.7 | 3.3 | -2.9 | -3.2 | -2.7 | 0/0/0 | 3.2 | 3.2 | 646.3 |
| 14 | 5.4 | 5.4 | 5.4 | -2.2 | -2.2 | -2.2 | 0/0/0 | 2.9 | 3.2 | 702.5 |
| 15 | 7.3 | 7.1 | 7.5 | -0.9 | -1.1 | -0.8 | 0/0/0 | 2.6 | 2.6 | 712.1 |
| 16 | 3.7 | 3.6 | 3.8 | -2.3 | -2.4 | -2.2 | 0/0/0 | 3.4 | 3.6 | 652.4 |

Positive Lewy body disease biomarkers are highlited with underbars.

**Figure S1.**
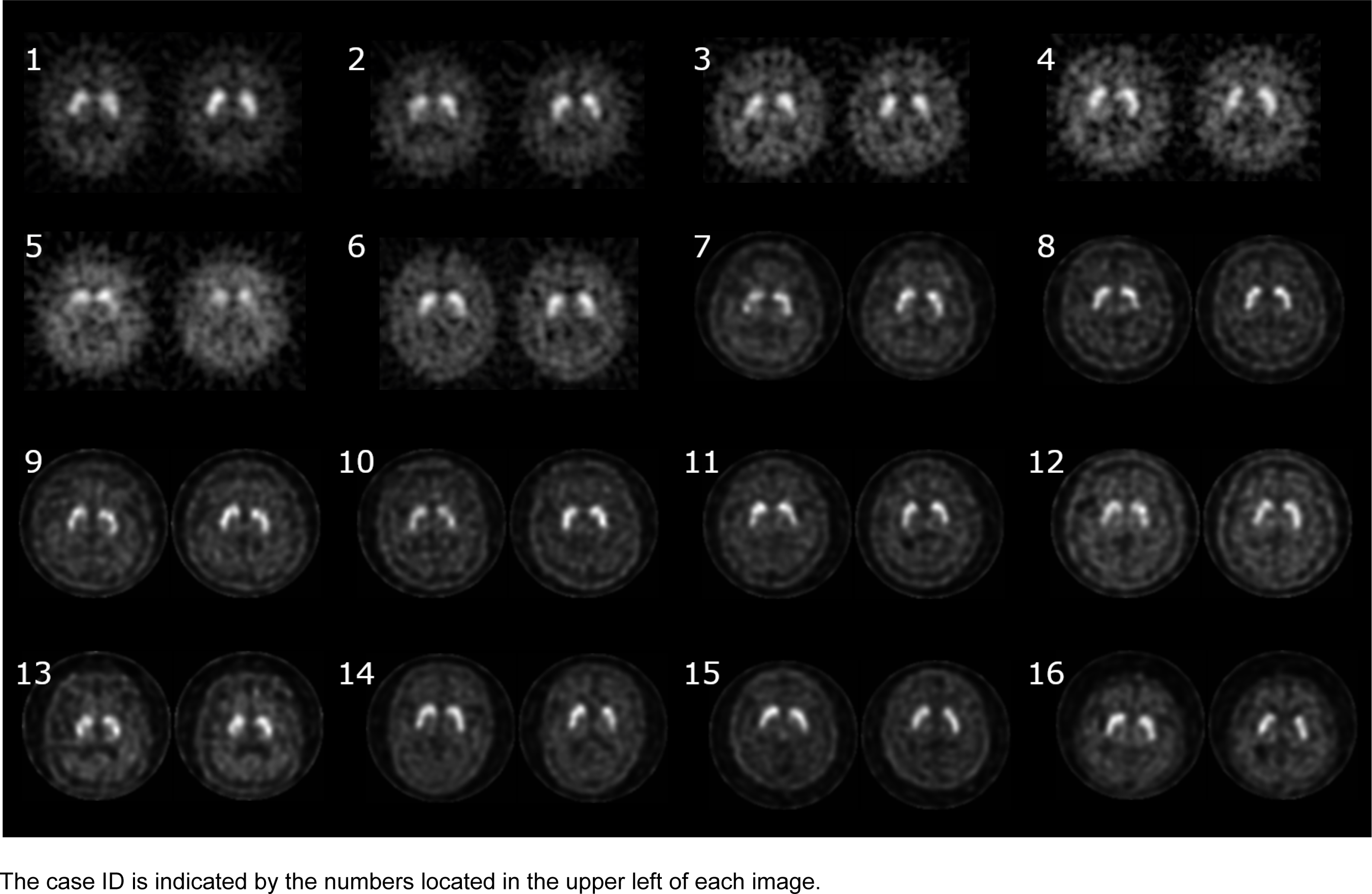
DAT-SPECT images of individual participants.

**Figure S2.**
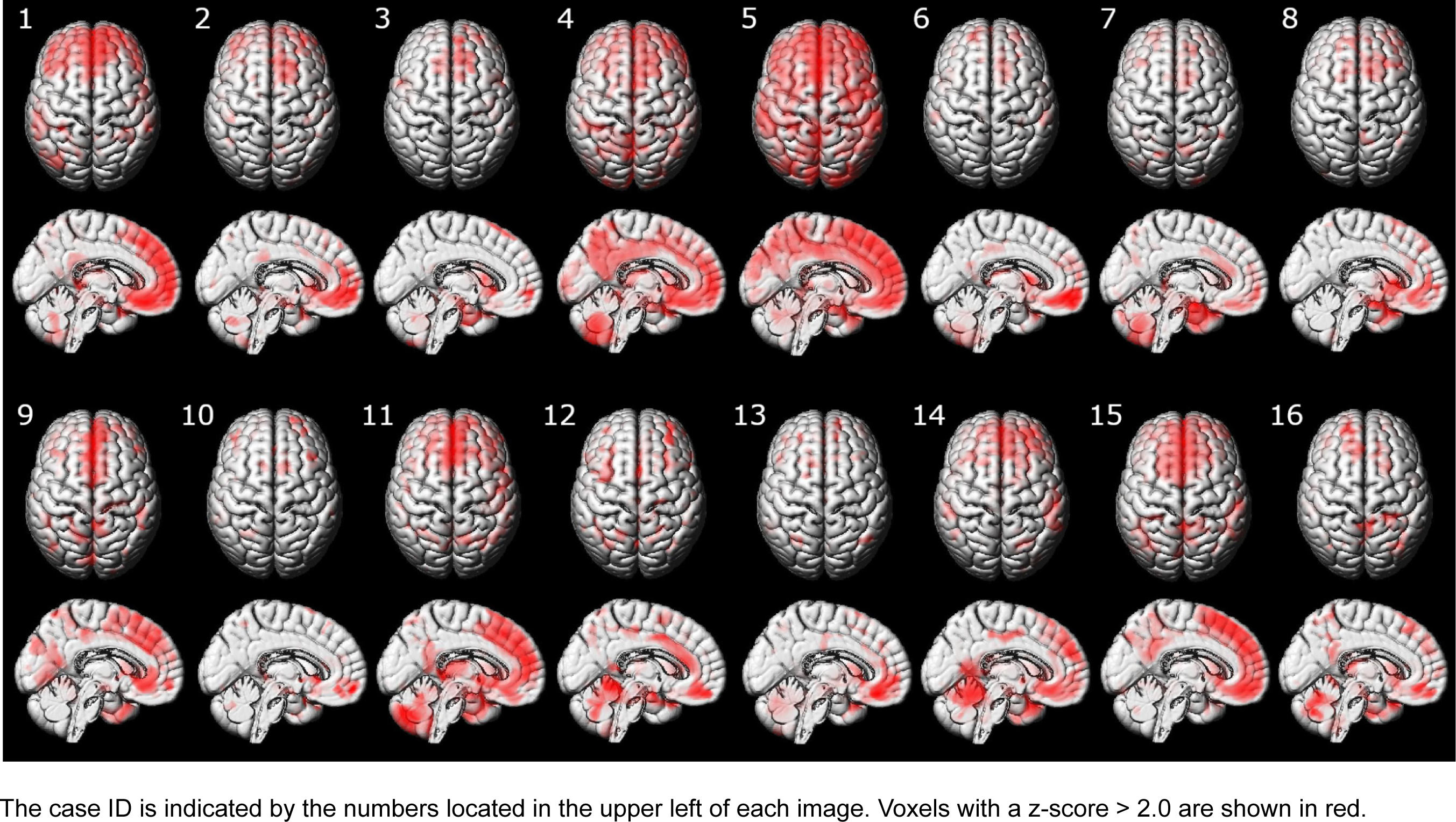
MRI voxel-based morphometry results of individual participants.

**Figure S3.**
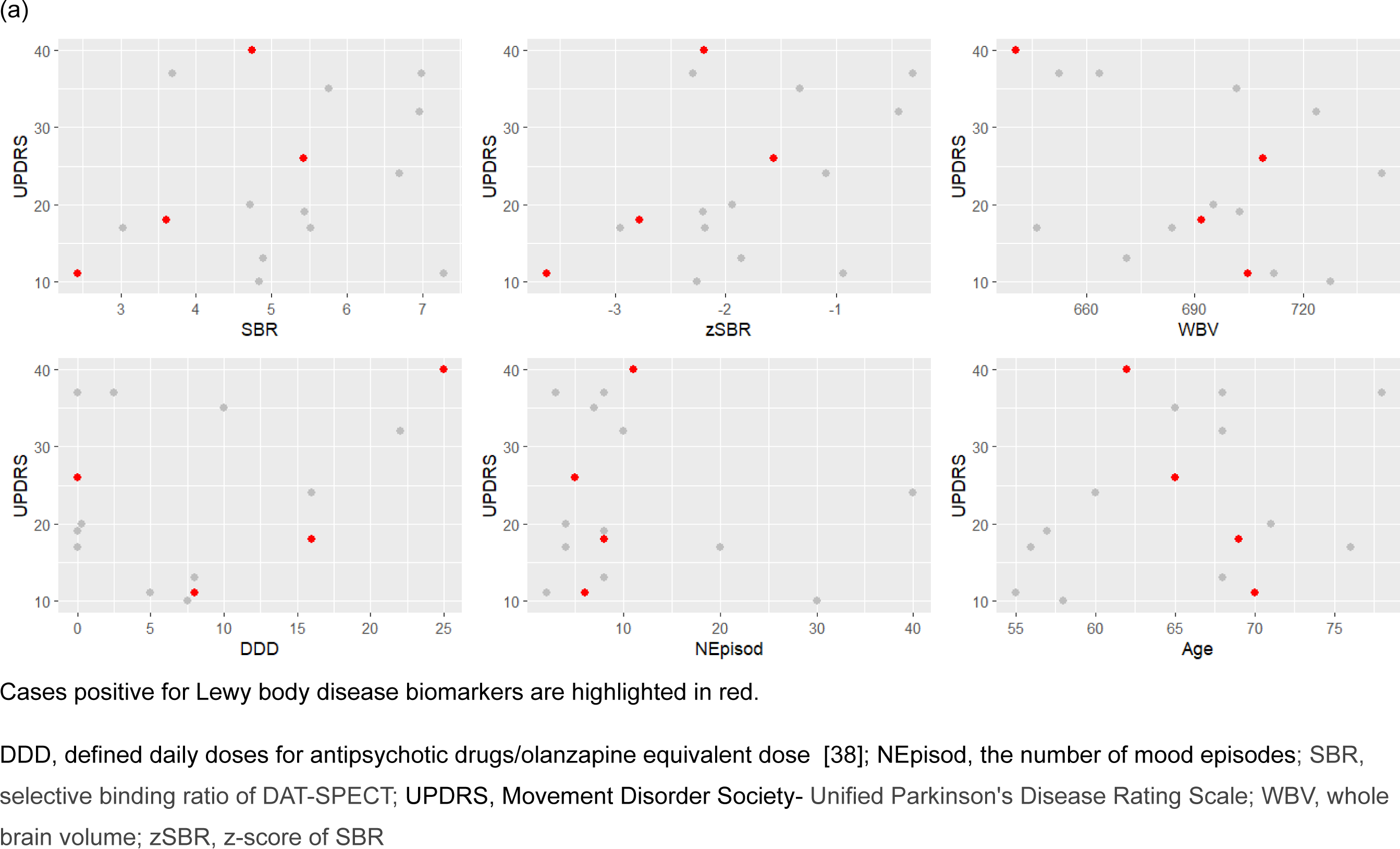
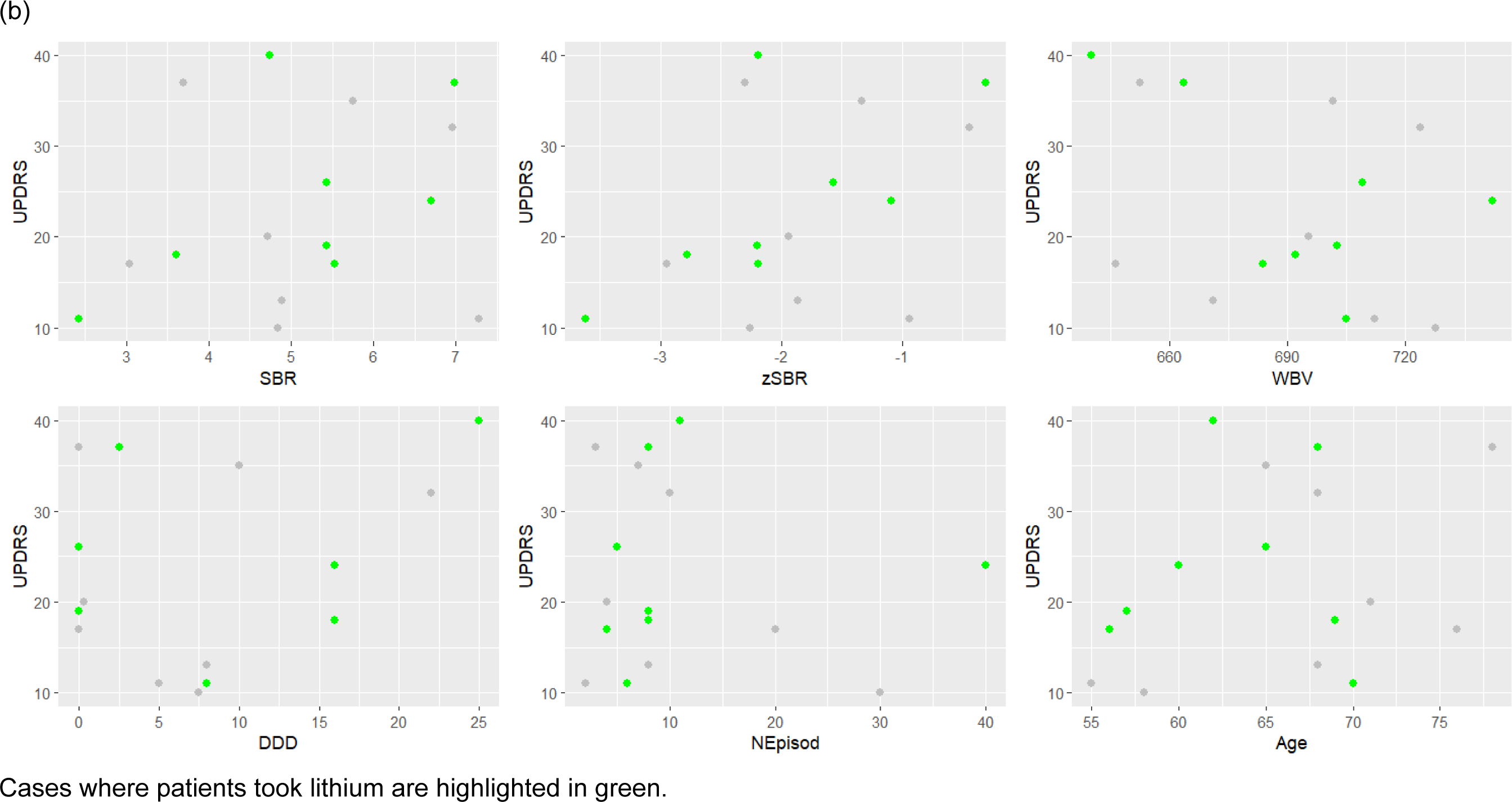
Brain-behavior correlations.

